# Evaluation of silver nanoparticles for the prevention of SARS-CoV-2 infection in health workers: *in vitro* and *in vivo*

**DOI:** 10.1101/2021.05.20.21256197

**Authors:** Horacio Almanza-Reyes, Sandra Moreno, Ismael Plascencia-López, Martha Alvarado-Vera, Leslie Patrón-Romero, Belén Borrego, Alberto Reyes-Escamilla, Daniel Valencia-Manzo, Alejandro Brun, Alexey Pestryakov, Nina Bogdanchikova

## Abstract

SARS-CoV-2 infection in hospital areas is of a particular concern, since the close interaction between health care personnel and patients diagnosed with COVID-19, which allows virus to be easily spread between them and subsequently to their families and communities. Preventing SARS-CoV-2 infection among healthcare personnel is essential to reduce the frequency of infections and outbreaks during the pandemic considering that they work in high-risk areas. In this research, silver nanoparticles (AgNPs) were tested *in vitro* and shown to have an inhibitory effect on SARS-CoV-2 infection in cultured cells. Subsequently, we assess the effects of mouthwash and nose rinse with ARGOVIT^®^ silver nanoparticles (AgNPs), in the prevention of SARS-CoV-2 contagion in health workers consider as high-risk group of acquiring the infection in the General Tijuana Hospital, Mexico, a hospital for the exclusive recruitment of patients diagnosed with COVID-19. We present a prospective randomized study of 231 participants that was carried out for 9 weeks (during the declaration of a pandemic). The “experimental” group was instructed to do mouthwash and nose rinse with the AgNPs solution; the “control” group was instructed to do mouthwashes and nose rinse in a conventional way. The incidence of SARS-CoV-2 infection was significantly lower in the “experimental” group (two participants of 114, 1.8%) compared to the “control” group (thirty-three participants of 117, 28.2%), with a 84.8% efficiency. We conclude that the mouth and nasal rinse with AgNPs helps in the prevention of SARS-CoV-2 infection in health personnel who are exposed to patients diagnosed with COVID-19.

## Introduction

On March 11, 2020, the World Health Organization declared the coronavirus disease 2019 (COVID-19) a worldwide public health problem; the responsible agent was named the severe acute respiratory syndrome coronavirus-2 (SARS-CoV-2) [1]. An estimated of 132,730,691 infected cases and 2,880,726 deaths have being reported as of April 8^th^, 2021 worldwide [2]. The infection triggered by SARS-CoV-2 can cause symptoms that appear from 2 to 14 days after exposure to the virus [3]. Caring for SARS-CoV-2 patients is a high risk exposition and causes complications due to the high mortality and morbidity [4]. In turn, there is a risk of nosocomial infections among patients with COVID-19 and patients without a diagnosis of atypical pneumonia [5]. To reduce the morbidity and morbidity rate of healthcare personal, a variety of public health interventions have been implemented, such as the adaptation of areas with negative air pressure, isolation areas for patients with COVID-19, and the mandatory use of personal protective equipment (gloves, goggles, overalls and N95/FFP2 respirators among others) to avoid contact with the respiratory and conjunctive tract [6]. International strategies focus on containment, diagnosis /monitoring, drug production and vaccines development against SARS-CoV-2 [7,8]. Different candidate drugs have been identified as antivirals against SARS-CoV-2 [9], but there are only very few drugs approved by the US Food and Drug Administration (FDA||) [10]. Their efficacy and safety are still under evaluation. The main strategy to prevent SARS-CoV-2 infection would be vaccination. So far, a dozen vaccines have been approved by different international organizations [11–15], but global access to vaccines is one of the drawbacks, also vaccination strategies can take years to be an effective solution. On the other hand, no vaccine has been created so far against any coronavirus causing the last three pandemics such as SARS-CoV in 2002 [16], MERS-CoV in 2012[17] and SARS-CoV-2 in 2019 [18]. Classic or traditional (attenuated or inactivated) vaccines have not worked because an Antibody-dependent enhancement (ADE) occurs [19]. Antibodies produced with one specific coronavirus subtype work, but they are not necessarily neutralizing, but rather opsonizing, leading to increased viral infectivity [20]. This leads to the fact that ADE-mediated secondary heterotypic infections caused by different coronavirus subtypes is the greatest risk factor [21,22]. SARS-CoV-2 infection in hospital areas is especially problematic, since the close interaction between health care personnel and patients diagnosed with COVID-19 which allows virus to be easily spread between them and subsequently to their families and communities. On the other hand, in previous research, the *in vitro* antiviral effect of AgNPs has been demonstrated [23,24]. Here, we confirmed the inhibitory effect of AgNPs in SARS-CoV-2 replication in cultured cells. For these reasons, a non-pharmaceutical public health intervention consisting of mouthwash and nose rinse with an AgNPs solution is proposed in order to reduce morbidity among healthcare personnel exposed to SARS-CoV-2 virus. We designed a prospective randomized controlled study of two-group (experimental vs. control) to evaluate the efficacy of mouthwash and nasal rinse with AgNPs solution for preventing SARS-CoV-2 infection in the health personnel at the General Tijuana Hospital in Mexico who works in high-risk areas with direct contact with patients infected and diagnosed with COVID-19.

## Materials and Methods

### Virus and cells

All *in vitro* studies were performed using the SARS-CoV-2 NL/2020 strain (BetaCoV/Netherlands/01) provided by the European Virus Archive global (EVAg). VeroE6 cells (ATCC Cat.No. CRL-1586), obtained from the cell repository at Centro de Investigación en Sanidad Animal, (CISA-INIA, Valdeolmos, Spain) were grown in Dulbecco’s modified Eagle’s medium (DMEM) supplemented with 10% fetal bovine serum (FBS), and L-glutamine (2 mM), penicillin (100 U/ml) and streptomycin (100 μg/ml), in a humid atmosphere of 5% CO_2_ at 37°C.

### Cell viability assays

Vero E6 cells were seeded in 96-well plates, and 24 hours later, when 80% confluence was reached, Argovit was added to the medium at serial two-fold dilutions, in triplicates. Vero E6 cells viability was tested in a long exposure experiment: Argovit (starting from a 1/2 dilution) was kept in the culture medium for 24, 48 and 72 hours. The treated cells were maintained at 37ºC in a 5% CO_2_ incubator and daily checked at the microscope. Viability of Vero E6 cells after treatment at the indicated times was checked using the MTS Cell Proliferation Assay (Promega) following manufacturer’s instructions. Viability percentages were calculated as the ratio OD_450_ nm treated wells/ OD_450_ nm Argovit-free wells x 100. The ODs corresponded to the mean of three replicas and were corrected by the corresponding blank wells without cells.

### *In vitro* infection experiments

Confluent monolayers of Vero E6 cells seeded on 12-well plates were infected with SARS-Cov2 at a multiplicity of infection (moi) of 0.001 plaque-forming units (pfu) per well in presence of metallic silver at serial two-fold dilutions from ½to 1/2048 (concentrations of 0.5% to 0.0004%). Plates were maintained at 37°C for three days. At 72 hpi supernatants were collected and titrated in Vero E6 cells grown in 12-well plates. After 45 minutes of adsorption, the inoculum was removed and semi-solid medium with 1% carboxymethyl-cellulose in 1X DMEM 5% FBS was added. Plates were incubated for 72 hours, then fixed and crystal violet stained. Plaques were counted and the percentage of infectivity for each Argovit dilution was calculated as follows: [1 – (number of plaques at the corresponding Argovit dilution)/ number of plaques with medium alone] x 100. Controls corresponding to 100% infectivity (infection in the absence of Argovit) as well as 100% viability (non-infected cells in medium containing Argovit) were also included. IC_50_ values were calculated using GraphPad software (Prism).

### Ethical considerations

The *in vivo* study was approved by the Research Ethics Committee of General Tijuana Hospital of the Institute of Public Health Services of the State of Baja California with favorable opinion number CONBIOETICA-02-CEI-001-20170 and was conducted in accordance with the Declaration of Helsinki. Signed consent was obtained from all participants. The objective of the study, the methodology and details of the experimental procedures and voluntary character of participation (including the right to withdraw from the study at any moment) were explained to all the participants. Participants were also informed on the anonymity and confidentiality of the data they reported during the study.

## Materials

The hygiene product for mouthwash and nasal rinse ARGOVIT^®^ AgNPs from the Investigation and Production Center Vector-Vita Ltd., made in Novosibirsk, Russia was applied. Metallic silver, polyvinylpyrrolidone, hydrolyzed collagen and distilled water concentrations in this solution are 0.06, 0.63, 0.31, and 99 wt.%, respectively. ARGOVIT^®^ is registered in Russia as an oral and nasal hygiene product since 2015.

### Study design

The *in vivo* study was carried out in the areas converted to care for patients diagnosed with atypical pneumonia and/or COVID-19 at General Tijuana Hospital belonging to the Mexican Ministry of Health. A 2-group randomized parallel study was conducted to compare the efficacy of mouthwash and nose rinse with the AgNPs 1 wt. % (0.6 mg of metallic Ag per mL) or direct spray to the oral cavity (experimental group); the second group was instructed to do mouthwash and nose rinse in a conventional way (control group). The investigation lasted 63 days (9 weeks) in the COVID-19 pandemic period, beginning in April 7 through June 9, 2020.

### Participants and environment

For the *in vivo* study, 231 volunteers were selected from General Hospital Tijuana. Men and women were included among the participants, ranging from 18 to 65 years of age, and various occupations. Personnel excluded from the study included persons with history of hypersensitivity to silver (rashes and other contraindications), a history of SARS-CoV-2 infection in the three months prior to the start of the study, any respiratory distress, and refusal to sign the informed consent.

### Instruments

The participants in the *in vivo* study were briefed on the objective of the study and were instructed on how to carry out the mouthwash and nose rinse. Subsequently, they were asked to complete a questionnaire where first identified the general data on the participants (Table 1.) and the second part, the participants reported weekly data related to their activities pertaining the study and their state of health (Table 2.). Experimental group were questioned about the presence of adverse effects by mouthwash and nose rinse. The diagnosis of COVID-19 was made by monitoring the signs and symptoms of the participants to finally confirm the infection status by RT-PCR (SuperScript III Platinum, Invitrogen, EEUU; Integrated DNA Technologies, Coralville, EEUU). Additionality, the two study groups were randomly selected for CT (Toshiba Aquilion 16, Japan) chest scan, RT-PCR analysis and clinical evaluation, to confirm the diagnosis of COVID-19. Participants with confirmed COVID-19 diagnosis were granted sick leave or were hospitalized.

**Table 1.**
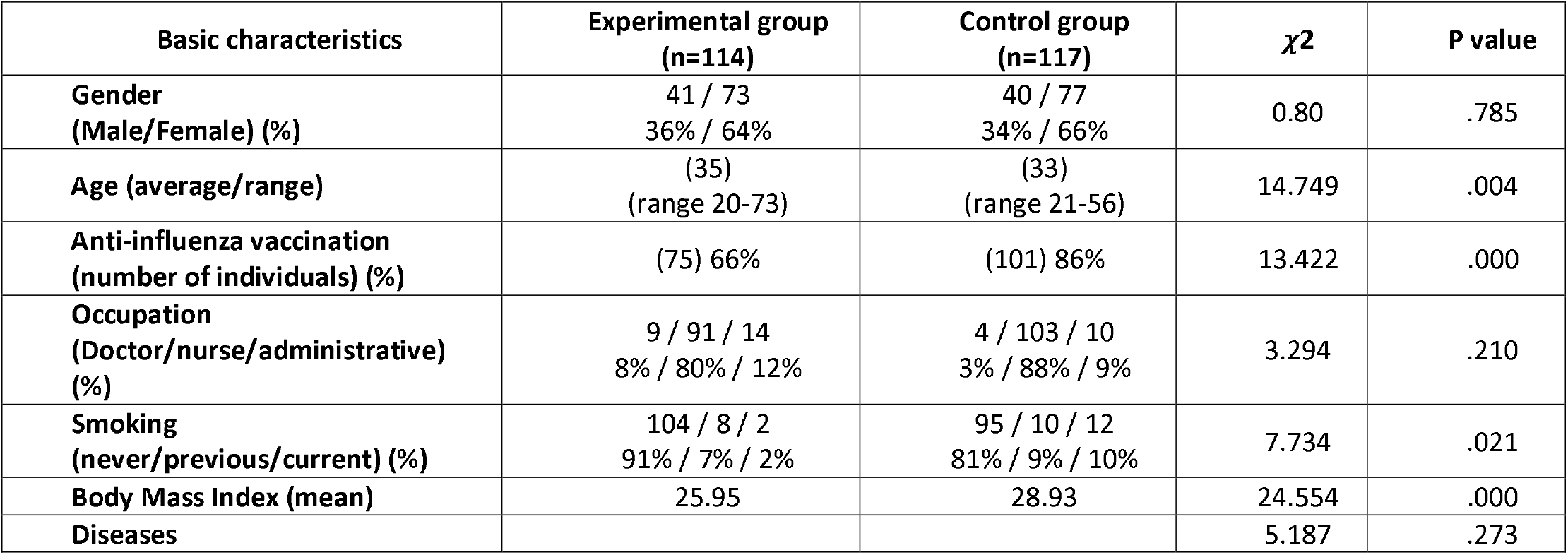
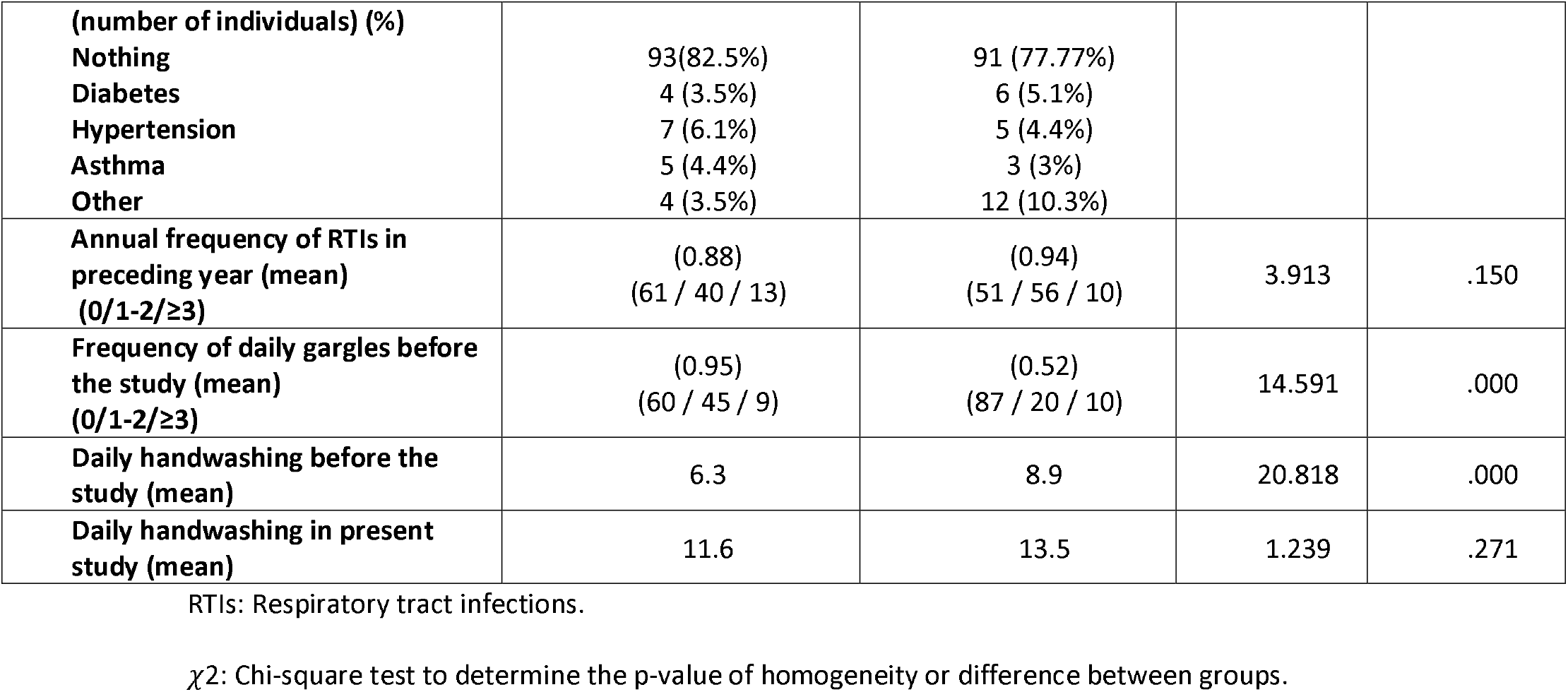
Characteristics of the experimental and control group.

**Table 2.** Prevention of SARS-CoV-2 infection in health workers.

| Variables | Experimental group (n=114) | Control group (n=117) | $\chi^2$ | P value |
| --- | --- | --- | --- | --- |
| Infected cases SARS-CoV-2 (number of individuals) (%) | 2 (1.8%) | 33 (28.2%) | 31.423 | 0.000 |
| Participants that did mouthwashes by gargle (number of individuals) (%) | 28 (24.56%) | 28 (23.93%) | 0.013 | 0.838 |
| Participants that did mouthwashes by spray (number of individuals) (%) | 34 (29.82%) | - | - | - |
| Participants that did mouthwashes by gargle and spray (number of individuals) (%) | 52 (45.61%) | - | - | - |
| Participants that did nasal rinses (number of individuals) (%) | 64 (54.70%) | 21(17.95%) | 36.18 | 0.00 |
| Number of daily gargles with AgNPs (mean) | 2 | 2.14 | 0.1667 | 0.563 |
| Number of mouthwashes with AgNPs spray (mean) | 2 | - | - | - |
| Number of mouthwashes by gargles and spray (mean) | 2 and 4 | - | - | - |
| Number of nasal rinses (mean) | 0.70 | 0.25 | 5.031 | 0.00 |
| Adverse reactions reported from using mouthwash with silver nanoparticles | 0 | - | - | - |
| Number of weekly contacts with patients COVID-19 (range) (mean) | (0-860) (169) | (0-729) (146) | - | - |
| Symptoms of RTIs (number of individuals) (%) | 21 (18.4%) | 42 (35.8%) | 8.891 | .003 |
| Occupation of infected individuals (doctor/nurse/administrative) | 0/2/0 | 2/29/2 | .515 | .773 |

AgNPs: silver nanoparticles.
RTIs: Respiratory tract infections.
$\chi^2$ : Chi-square test to determine the p-value.

### Randomization and experimental intervention

Eligible participants for the *in vivo* study were randomized using a computer generated block scheme and stratified according to duty position, work shifts and the area/department of the service at General Tijuana Hospital. Individuals from experimental group were provided with a 50 mL spray bottle containing AgNPs solution with 1 wt% concentration (0.6 mg/mL metallic silver). They were instructed to mix 4 to 6 spray shots (corresponding to volume ∼ 0.5 mL) of this solution with 20 mL of water and to gargle with obtained solution for 15 to 30 seconds at least 3 times a day, also nasal lavages on the inner part of the nasal alae and nasal passage with the same solution using a cotton swap twice a day. As a second option, they were instructed to cover evenly the oral cavity with the direct 1 to 2 spray shots of solution without its previous dilution in water. Participants of the control group were instructed to do mouthwash and nose rinse with a conventional mouthwash the way they normally did before the study.

### Data analysis

Data analysis was performed through the SPSS statistical program (version 26). Descriptive analysis was done with frequencies, percentages, means and ranges. To check the statistical difference between the results of the experimental and control groups, the Pearson’s “Chi-Square” tests, the “Fisher’s exact test” and the “likelihood ratio” were used. The symmetry measures for 2 x 2 contingency tables were determined by the Statistics of “Phi”, “Contingency Coefficient” and “Cramer’s V”. A logistic regression was performed to corroborate the efficacy of performing mouthwash and nose rinse with AgNPs and its impact on the incidence of SARS-CoV-2 infections among the participants under similar conditions. Likewise, the Chi-Square statistic was applied to determine the p-value in order to determine the homogeneity or heterogeneity between the experimental group and the control group.

## Results

### Antiviral activity of AgNPs in Vero E6 cell cultures

To determine the efficacy of AgNPs against SARS-CoV-2 *in vitro*, we first analyzed its cytotoxicity on cultured Vero E6 cells. Cells were seeded in 96-iwell plates and two-fold serial dilutions of AgNPs added to the medium, in triplicates. The cells were then incubated for 24, 48 or 72 hours. Cells grown in the presence of AgNPs over 24 hours showed the highest viability values over 1/32 dilution (or 0.03% of AgNPs) (Figure 1A). In order to ensure viral infection, further experiments were performed with [AgNPs] ≤ 0.03%. To analyze the effect of AgNPs on virus infectivity Vero E6 cells were infected with a fixed amount of virus and different concentrations of AgNPs, starting at 0.03%, were added to cells. At 72 hours post-infection (pi) supernatants were collected and titrated in order to determine virus yields normalized to those reached in medium alone (Figure 1B). Although AgNPs did not totally abolish viral production, infection was clearly controlled to some extent with a reduction of about 80% at a concentration of 0.03%. A 50% inhibitory concentration was determined by curve fitting (non-linear regression) (Fig. 1BC).

**Fig. 1.**
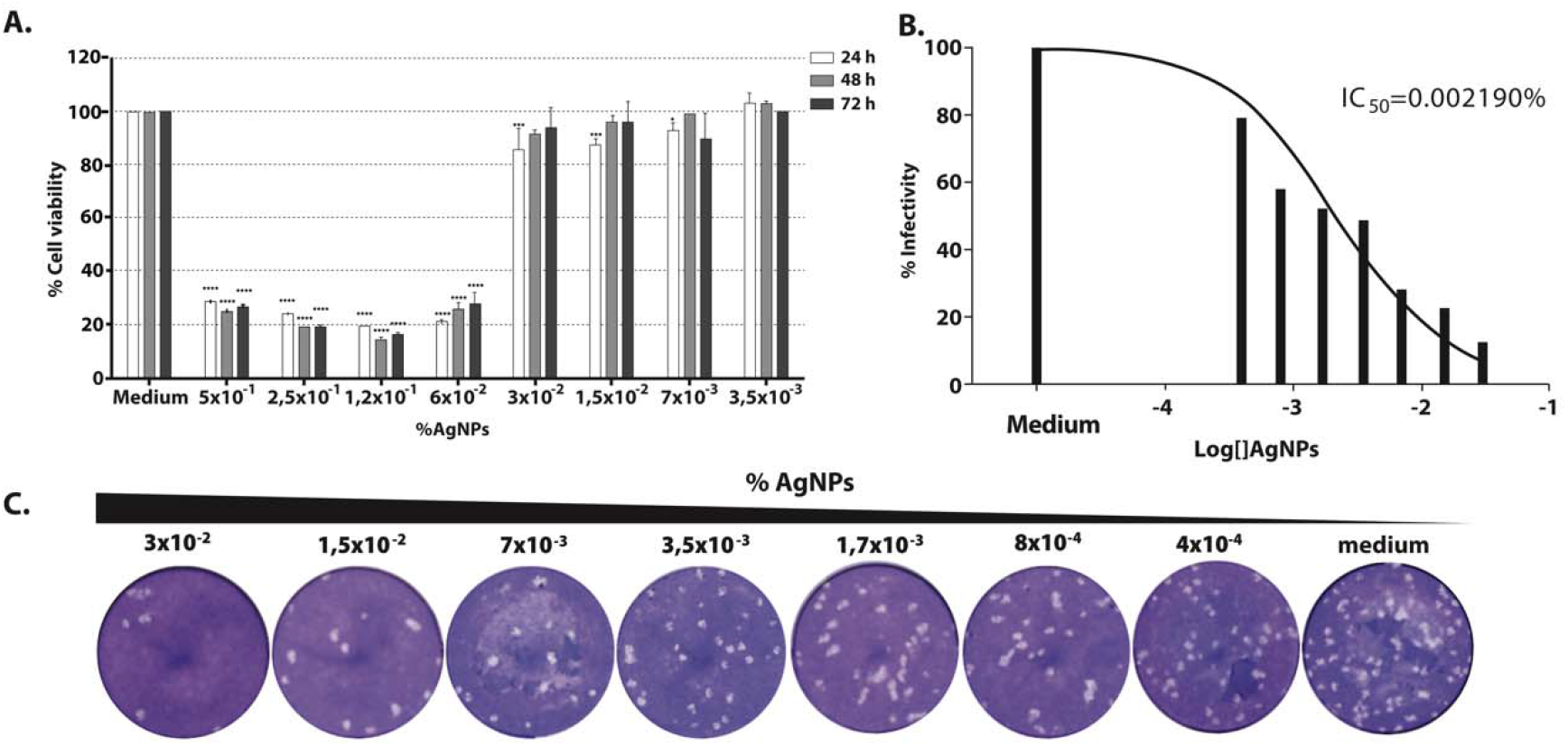
Antiviral activity of AgNPs in Vero E6 cell cultures. **A**. Effect of AgNPs on viability of cultured Vero cells. Serial two-fold dilutions of AgNPs were added in triplicate to the medium of Vero cells seeded in 96-well plates. Viability at 24 (white bars), 48 (grey bars) and 72 h (black bars) was checked by the MTS Cell Proliferation Assay (Promega) and calculated as described in Methods. Statistically significant differences when compared to the corresponding 100% of cell viability (by 2 way ANOVA) are indicated. *, p < 0.05; ***, p < 0.001; ****, p < 0.0001. **B**. Percentages of infectivity values for each AgNPs concentration. The values were normalized to those in the absence of Argovit (medium, 100%) and fitted using a non-linear regression algorithm. IC_50_: 50% inhibitory concentration. **C**. Representative SARS-CoV-2 plaque formation in presence of different dilutions of AgNPs.

### Description of the groups

From the total of 231 participants, 114 were assigned to the experimental group, including 41 men (36%) and 73 women (64%), with an average age of 35 years. The control group included 117 participants, 40 men (34%) and 77 (66%) women with an average age of 33 years. 66% of the participants in the experimental group were vaccinated against influenza virus during the last year, while in the control group the vaccinated participants accounted for 86%. Regarding the profession of health personnel, there were 8% doctors, 80% nurses and 12% administrative in the experimental group and 3% doctors, 88% nurses and 9% administrative in the control group. A reduced number of participants reported being smokers (9% in the experimental group vs. 19% in the control group); as well as not having chronic diseases such as: diabetes (3% in the experimental group vs. 5% in the control group); hypertension (6% in the experimental group vs. 4% in the control group); and asthma (4% in the experimental group vs 3% in the control group). Most participants reported becoming ill with respiratory tract infections ≤ 2 times in the past year (89% in the experimental group and 91% in the control group). The frequency of daily handwashing and mouthwash prior to the study, in the experimental group were: 6.3 handwashing and 0.95 average daily mouthwash, and in the control group 8.9 handwashing and 0.52 average daily mouthwash. Studied groups characteristics and Chi Square tests results which determine the significance of differences between groups are presented in Table 1.

## Results analysis

The incidence of SARS-CoV-2 infection (p = 0.000), was significantly lower in the experimental group vs the control group, where 1.8% (2 participants out of 114) and 28.2% (33 participants out of 117) were infected respectively (Table 2); frequency of mouthwashes (gargles and/or spray application) and nasal application daily with AgNPs, performed only by the experimental group, on average 2 mouthwashes made by 28 participants who only did it in the form of gargles, 2 daily applications by 34 participants who only used it in spray; while the 52 participants who chose to do both types of rinses, performed them on average 2 times as a gargle and 4 times as a spray application; on average 0.70 nasal rinse made by 64 participants; weekly number of contacts with patients diagnosed with COVID-19 or atypical pneumonia during the study period: (0 - 860, 169 on average) in the experimental group and (0 to 729, 146 on average) in the control group; percentage of individuals who presented symptoms of respiratory tract infection (RTI’s) at some point during the study; 18.4% in the experimental group and 35.8% in the control group; no adverse reactions were reported by performing mouthwash and nose rinse with AgNPs.

Pearson’s Chi-Square tests, the “Fisher’s exact test” and the “likelihood ratio” were run to check the statistical difference between the experimental and control groups. If the use of AgNPs had not been effective and 1) the protection measures used in the General Tijuana Hospital were very effective, the same low contagion level (1.8%) would have been presented in both groups; 2) if the protection measures were not effective, the same high contagion level (28.2%) would have been presented in both groups. Results were significant at 99% reliability for both one-tailed and two-tailed tests verified the difference in group results. The predictive model based on the experimental data reports that the odds of a participant who is treated with the AgNPs are 22 times higher than those of a participant who is not treated. The logistic model predicts that if all participants had mouthwashes and nasal rinses, the probability of success (they did not become infected with SARS-CoV-2) would be 84.8%. Table 3 shows the good fit of the data to the model taking into account that the probability of non-contagion depends on whether or not performed AgNPs solution, but the number of mouthwashes and nasal rinse on average per day (gargling and nasal rinse, spray application or combined) was also taken into account.

**Table 3.**
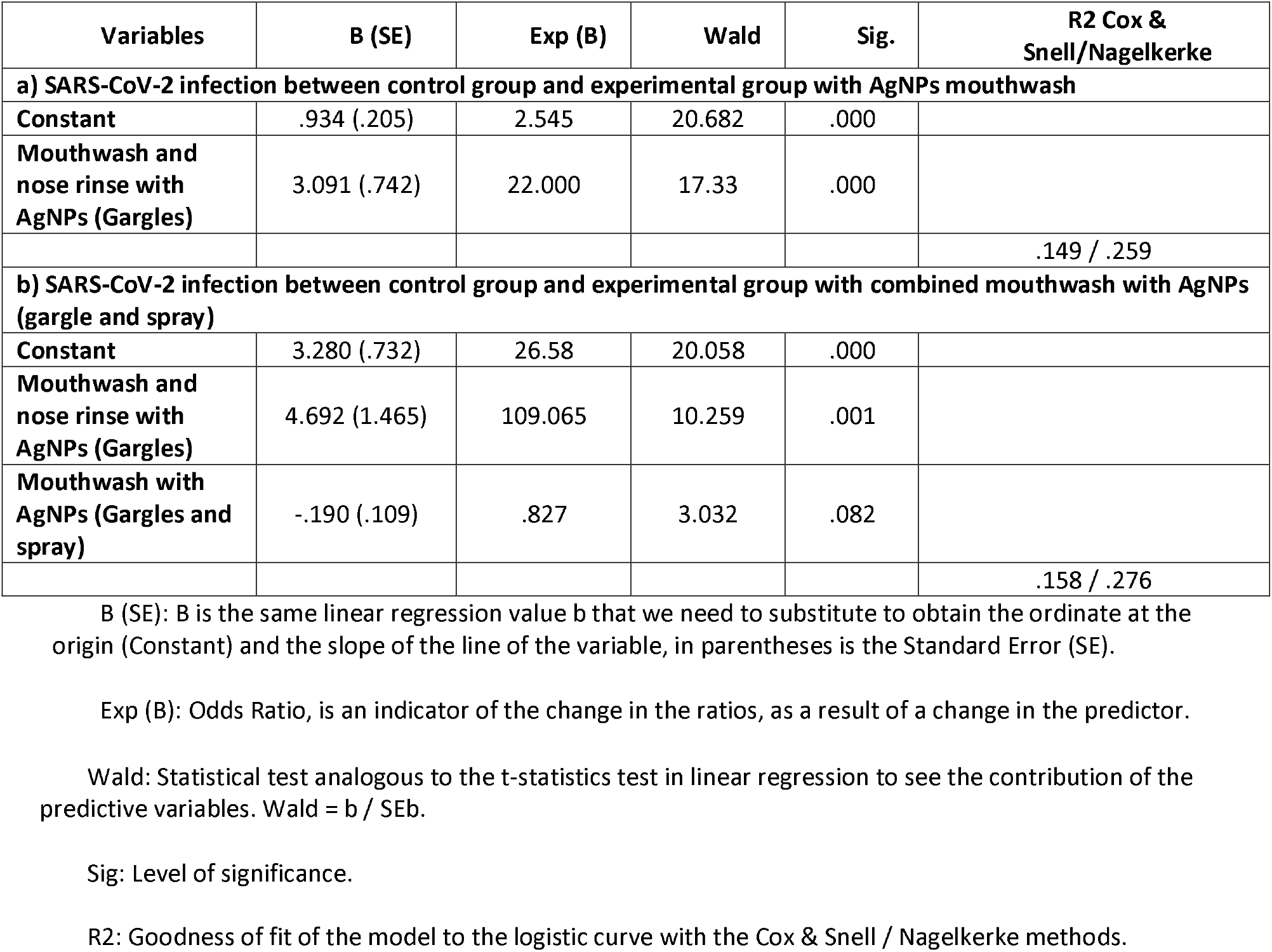
Results of the logistic regression of SARS-CoV-2 infection between control and experimental groups.

The dichotomous independent variable (participated in the experiment or not) is highly significant according to the value of the Wald statistic. The interesting thing about the logistic regression is the statistic Exp (B), which in this case tells us that for each person who “‘DOESN’T become infected” with SARS-CoV-2 in the control group, there are 22 people “NOT INFECTED” when they perform mouthwash and nose rinses with AgNPs in the experimental group in similar environments or under similar conditions. To find out whether the number of times mouthwashes and nasal rinse is performed with AgNPs per day has an effect, the quantitative variable was added to the dichotomous variables of the previous model where the average number of mouthwashes and nasal rinse was combined as in our sample. It was 2 gargle mouthwashes, 1 nasal rinse, and 2 spray mouthwashes. When we assume that 4 mouthwashes were performed daily (2 gargle and 2 spray) and 1 nasal rinse, the model improves with respect to only saying that the person participated in the experiment without considering the number of applications. The EXP (B) Statistic rises from 22 to 109 when considering mouthwash in both gargles and spray. That is, for each person who becomes infected with SARS-CoV-2 by performing 4 mouthwashes (2 gargles and 2 spray) and 1 nasal rinse, per day 109 people will not be infected.

## Discussion

Silver nanoparticles are known to have a powerful antimicrobial effect and there have been several studies that also demonstrated their antiviral effect [25]. The mechanism by which AgNPs interact with viruses is not yet clear, but it has been proposed that AgNPs, as well as other metallic nanoparticles, interfere with the structural proteins of the virus by inhibiting their ability to bind with cell receptors or bind to genetic material of viruses by inhibiting their replication[26]. Since the nanoparticles used in these experiments are mixed with hydrolyzed collagen, a compound that stabilizes nanoparticles and reduces its toxicity in cell culture [28], we were able to assess higher concentrations than those used in other articles. Jeremiah et al. showed that in Vero E6 cells AgNPs alone are toxic at concentrations of 100 ppm. In contrast, we observed that collagen AgNPs are not toxic up to concentrations of 0.03% (312 ppm). Despite this fact, the toxicity of silver nanoparticles in cell culture narrows the range of concentrations at which we can study therapeutic effects against viral infections *in vitro*. Taking this limitation into account we could observe a clear dose-dependent inhibitory effect over SARS-CoV-2 infectivity.

To our knowledge, this study is the first experimental trial where AgNPs are applied as a mouthwash and nose rinse solution for the prevention of SARS-CoV-2 infection in health care personnel working with COVID-19 diagnosed patients. The study was carried out in a prospective, controlled and randomized way for 9 weeks in the period of pandemic by COVID-19 from April to June 2020 in Tijuana Baja California, Mexico. The characterization of the two groups showed no significant statistical differences in gender, occupation and comorbidities related to the complication of SARS-CoV-2 infection. However, a statistically significant difference was identified in variables such as: age (p = 0.004), average age 35 in the experimental group and average age 33 in the control group; anti-influenza vaccination (p = 0.000), (66%) in the experimental group and (86%) in the control group, even though, vaccination against influenza virus reduces morbidity/mortality among medical personnel exclusively against influenza virus, but does not have any antigenic similarity with SARS-CoV-2 virus, as an example in this study, the control group reported a higher percentage of participants vaccinated against influenza and a higher percentage of infection by SARS-Cov2; Body Mass Index (p = 0.000), (25.95) in the experimental group and (28.93) in the control group, both groups classifying within the 25-29 Overweight level; frequency of daily gargles before the study (p = 0.000), (0.95); frequency of daily handwashing before the study (p = 0.000), (6.3) in the experimental group and (8.9) in the control group, the experimental group had the lowest number of COVID-19 infections, being the group that washed their hands less frequently, this indicates that AgNPs contributed to mitigating the SARS-CoV-2 infection in the experimental group, consistent handwashing has previously been recognized as a protective factor against SARS and influenza [29], in addition, most respiratory pathogens are spread by direct contact, droplets, and fomites [30]; the most important statistically significant difference, the incidence of SARS-CoV-2 infection was significantly lower in the experimental group (1.8%) than the control group (28.2%) (p=0.000); and finally, the report of symptoms related to the SARS-CoV-2 infection, there was a statistically significant difference (p = 0.003) reported by the experimental group (18.4%), while in the control group (35.8%).

These results corroborate the relevance of oral and nasal hygiene with AgNPs in the prevention of respiratory tract infection to minimize the risk of contagion by COVID-19 in health personnel. Scientific evidence supports preventive oral and nasal health regimens, including the use of mouthwashes, gargles, and nasal washes, as important components of SARS-CoV-2 infection control practices [31–33]. Using a logistic regression, the strong relationship between the fact of mouthwashes and nose rinse in the health personnel and the mitigation of the SARS-CoV-2 contagion with an efficiency of 84.8% is proved. The results provide clinical evidence to confirm the prevention of SARS-CoV-2 infection in health personnel who performed mouthwash and nose rinse solution with AgNPs. In the midst of the global pandemic, the results of *in vitro* infection experiments (effect of AgNPs on virus infectivity and determination of the 50% inhibitory concentration) and Argovit registration as an oral and nasal hygiene product in Russia since 2015, and with antimicrobial action for oral and nasal hygiene since 2019, were decisive to apply the AgNPs *in vivo* in the oral and nasal cavity to confirm its effects against SARS-CoV-2. The present study also showed that no harmful side effects were observed in the 114 participants who used AgNPs as a mouthwash and nose rinse solution for 9 weeks.

The relevance of mouthwash and nasal rinses as a prevention strategy for mitigating SARS-CoV-2 infections among health personnel also extends to other high-risk areas such as dental procedures, in these cases, specific protocols are suggested to avoid COVID-19 contamination. The oropharynx and nasopharynx are the initial entry sites where SARS-CoV-2 replicates and can produce a viral load of 1.2 × 10^8^ infectious copies / per mL [34]. The odontologist works with a high risk of exposure during routine dental procedures. In addition, the use of different devices that generate a high amount of aerosols (rotary instruments and ultrasounds), which can be inhaled through the respiratory route, these aerosols can also carry viral particles that come into contact with the conjunctiva and also cause infection, on the other hand these residues can be deposited on different surfaces, on the work clothes, masks, hands and clothes of the patient. Dental cabinets are considered a possible source of contamination and nosocomial infection [35]. Contamination by nasal excretions can carry thousands of infectious particles, since a higher viral load has been demonstrated in the nasal cavity compared to the oral cavity [36]. Recent reports indicate that total anosmia or partial loss of the sense of smell are early markers of SARS-CoV-2 infection. This phenomenon can be caused by the expression of the receptors of the transmembrane protein of the angiotensin converting enzyme II (ACE2) and by a type II transmembrane serine protease (TMPRSS2), responsible for the entry of the virus into nasopharyngeal cells and present direct damage secondary to SARS-CoV-2 viral replication in olfactory receptor neurons (ORNs) located in the olfactory epithelium [37]. These results justify making changes to the international recommendations [35] on new dental clinical practices and the prevention of SARS-CoV-2. Since the dental clinic procedure requires that the patient, after passing the sanitary filter, has to perform a mouth rinse (on average 30 minutes before the consultation), followed by a second mouth rinse when seated in the dental chair. At no time do international recommendations[35] speak of nasal washes, when it has been demonstrated that by nasal exhalation the spread of SARS-CoV-2 to health personnel can occur. The evidence presented in this study suggests that the application of mouthwashes and nose rinse can significantly reduce the viral load in these areas, to reduce transmission, in addition to the use of personal protective equipment by healthcare personnel.

The lack of efficient and specific SARS-CoV-2 antiviral therapies could be due in large part to the underuse of nanotechnologies [38], non-pharmaceutical therapies based on nanotechnology applications are needed as preventive measures, to mitigate the nosocomial transmission of SARS-CoV-2 among health personnel. We present a new intervention strategy that uses AgNPs as a mouthwash and nasal rinse solution that can reduce the spread, transmission and/or pathogenicity of SARS-CoV-2 associated with COVID-19. Due to the frequent generation of aerosols by direct treatment with patients with COVID-19, the associated risk of virus transmission was very low among health personnel who performed mouthwashes and nasal washes with AgNPs. The antiviral effect of mouthwashes and nose rinse with AgNPs can decrease the viral load in the oral and nasal cavity, as well as inhibit the proliferation of the virus and temporarily reduce the risk of transmission. These results were demonstrated in the clinical field of the health personnel of the Tijuana General Hospital.

## Conclusions

This prospective randomized study demonstrates that mouthwash and nose rinse with AgNPs is effective in decreasing SARS-CoV-2 infection rate. To our knowledge, this study is the first experimental *in vitro* e *in vivo* trial where AgNPs as mouthwash and nasal rinse solution are applied for SARS-CoV-2 contagion prevention. The results of this investigation are revealing and encouraging, due to the fact that the health personnel exposed to an excessive viral load due to the number of COVID-19 patients attended (169 patients attended on average per week per person) were not infected, which is attributed to rinsing with AgNPs. Proved its inhibitory effect on SARS-CoV-2 infectivity *in vitro* it is inferred that the use of AgNPs as a mouthwash and nose rinse will be very useful as a prophylactic for the prevention of SARS-CoV-2, not only for health care personnel, but also as additional protection for the general population.

## Supporting information

Supplemental Clinical Trial Registration Number

## Data Availability

Data are available by the corresponsal author, please contact via

## Acknowledgments

The authors of this study would like to thank the staff of General Tijuana Hospital for their invaluable participation in this research during the extremely difficult period of the COVID-19 pandemic. The health care personnel are under intense workout during this pandemic. The authors would also like to thank CONACyT “International Network of Bionanotechnology with an Impact on Biomedicine, Food and Biosafety” and Tomsk Polytechnic University Competitiveness Enhancement Program, project VIU-RSCBMT-197/2020 for their important collaboration in the field of fundamental and applied research on nanomaterials in nanomedicine.

