## Supplemental Clinical Trial Registration Number for "Evaluation of silver nanoparticles for the prevention of SARS-CoV-2 infection in health workers: *in vitro* and *in vivo*"

**ClinicalTrials.gov Protocol Registration and Results System (PRS) Receipt**
Release Date: May 18, 2021

**ClinicalTrials.gov ID: NCT04894409**

### Study Identification

Unique Protocol ID: CONBIOETICA-02-CEI-001-20170

Brief Title: Evaluation of Silver Nanoparticles for the Prevention of COVID-19

Official Title: Evaluation of Silver Nanoparticles as an Oropharyngeal Product (Mouthwash) and Nasal Hygiene, by Health Personnel Working at the Tijuana General Hospital Exposed to Patients Diagnosed With Atypical Pneumonia Caused by SARS-CoV-2

Secondary IDs:

### Study Status

Record Verification: May 2021

Overall Status: Completed

Study Start: April 24, 2020 [Actual]

Primary Completion: June 30, 2020 [Actual]

Study Completion: September 29, 2020 [Actual]

### Sponsor/Collaborators

Sponsor: Cluster de Bioeconomia de Baja California, A.C

Responsible Party: Sponsor

Collaborators: Bionag SAPI de CV
General Hospital Tijuana

### Oversight

U.S. FDA-regulated Drug: No

U.S. FDA-regulated Device: No

Unapproved/Uncleared Device: No

U.S. FDA IND/IDE: No

Human Subjects Review: Board Status: Approved
Approval Number: CONBIOETICA-02-CEI-001-20170
Board Name: General Hospital Tijuana Research Ethics Committee
Board Affiliation: Institute Public Health Services of the State of Baja California

Address:

1819 Fresnillo Street, Madero, Zip Code 22040 Tijuana, Baja California, México.

Data Monitoring: No

FDA Regulated Intervention: No

### Study Description

Brief Summary: In this research, silver nanoparticles (AgNPs) were tested in vitro and shown to have an inhibitory effect on SARS-CoV-2 infection in cultured cells. Subsequently, the investigators assessed the effects of mouthwash and nose rinse with ARGOVIT® silver nanoparticles (AgNPs), in the prevention of SARS-CoV-2 contagion in health workers consider as high-risk group of acquiring the infection in the General Tijuana Hospital, Mexico, a hospital for the exclusive recruitment of patients diagnosed with COVID-19.

Detailed Description: SARS-CoV-2 infection in hospital areas is of a particular concern, since the close interaction between health care personnel and patients diagnosed with COVID-19, which allows virus to be easily spread between them and subsequently to their families and communities. Preventing SARS-CoV-2 infection among healthcare personnel is essential to reduce the frequency of infections and outbreaks during the pandemic. In a first step, silver nanoparticles (AgNPs) were tested in vitro to determine an inhibitory effect on SARS-CoV-2 infection in cultured cells. Subsequently, the investigators assess the effects of mouthwash and nose rinse with ARGOVIT® silver nanoparticles (AgNPs), in the prevention of SARS-CoV-2 contagion in health workers consider as high-risk group of acquiring the infection in the General Tijuana Hospital, Mexico, a hospital for the exclusive recruitment of patients diagnosed with COVID-19. The investigators present a prospective randomized study of 231 participants that was carried out for 9 weeks (during the declaration of a pandemic). The "experimental" group was instructed to do mouthwash and nose rinse with the AgNPs solution; the "control" group was instructed to do mouthwashes and nose rinse in a conventional way.

### Conditions

Conditions: Coronavirus Disease 2019 (COVID-19)

Keywords: Silver nanoparticles
SARS-CoV-2
COVID-19
Infection control

### Study Design

Study Type: Interventional

Primary Purpose: Other

Study Phase: N/A

Interventional Study Model: Crossover Assignment
Eligible participants for the in vivo study were randomized using a computer generated block scheme and stratified according to duty position, work shifts and the area/department of the service at General Tijuana Hospital. Individuals from experimental group were provided with a 50 mL spray bottle containing AgNPs solution with 1 wt% concentration (0.6 mg/mL metallic silver). Participants were instructed to mix 4 to 6 spray shots (corresponding to volume ~ 0.5 mL) of this solution with 20 mL of water and to gargle with obtained

solution for 15 to 30 seconds at least 3 times a day, also nasal lavages on the inner part of the nasal alae and nasal passage with the same solution using a cotton swap twice a day. As a second option, participants were instructed to cover evenly the oral cavity with the direct 1 to 2 spray shots of solution without its previous dilution in water.

Number of Arms: 2

Masking: None (Open Label)

Allocation: Randomized

Enrollment: 231 [Actual]

### Arms and Interventions

| Arms | Assigned Interventions |
| --- | --- |
| Experimental: Experimental group<br>The experimental group was instructed to do mouthwash and nose rinse with the AgNPs solution. | Device: Mouthwash and nose rinse with the AgNPs solution<br>The "experimental" group was instructed to do mouthwash and nose rinse with the AgNPs solution for the prevention of SARS-CoV-2 infection in health workers |
| Active Comparator: Control group<br>The "control" group was instructed to do mouthwashes and nose rinse in a conventional way. | Device: Mouthwashes and nose rinse in a conventional way<br>The control group was instructed to do mouthwashes and nose rinse in a conventional way |

### Outcome Measures

Primary Outcome Measure:

1. Incidence of SARS-CoV-2 infection in the experimental group.
   Percentage of participants infected of SARS-CoV-2 in the experimental group.

   [Time Frame: 9 weeks]
2. Incidence of SARS-CoV-2 infection in the control group.
   Percentage of participants infected of SARS-CoV-2 in the control group.

   [Time Frame: 9 weeks]

Secondary Outcome Measure:

3. Number of participants with adverse reactions by AgNPs.
   Number of participants with adverse reactions by performing mouthwash and nose rinse with AgNPs.

   [Time Frame: 9 weeks]

### Eligibility

Minimum Age: 20 Years

Maximum Age: 73 Years

Sex: All

Gender Based: No

Accepts Healthy Volunteers: Yes

Criteria: Inclusion Criteria:

- Men and women health workers in the General Tijuana Hospital, Mexico who works in high-risk areas with direct contact with patients infected and diagnosed with COVID-19.

Exclusion Criteria:

- persons with history of hypersensitivity to silver (rashes and other contraindications),
- a history of SARS-CoV-2 infection in the three months prior to the start of the study, any respiratory distress,
- and refusal to sign the informed consent.

### Contacts/Locations

Central Contact Person: Horacio Almanza-Reyes, PhD


Central Contact Backup: Ismael Plascencia-López, PhD


Study Officials:

Locations: **Mexico**
Tijuana General Hospital
Tijuana, Baja California, Mexico, 22310
Contact: Alberto Reyes Escamilla, MD 52-664-684-0078 Ext. 2449
Contact: Daniel Valencia, Mtro 52-664-477-5316
Principal Investigator: Horacio Almanza-Reyes, PhD

### IPDSharing

Plan to Share IPD: No

Available IPD/Information:
